# Increased sensorimotor noise in Tourette syndrome

**DOI:** 10.1101/2025.01.03.24319737

**Authors:** Aikaterini Gialopsou, Mairi Houlgreave, Stephen R. Jackson

## Abstract

Tourette syndrome (TS) is a neurological disorder characterised by the occurrence of vocal and motor tics^1^. The pathophysiology of TS has been linked to a substantial reduction in the number of inhibitory GABAergic interneurons found within the striatum^2^, leading to increased neural ‘noise’ within the cortical-striatal-thalamic-cortical [CSTC] circuit implicated in movement production^3^. In the current study, we used electroencephalography (EEG) to investigate increased neural noise in a group of 19 adults with TS compared to a matched neurotypical control group. We operationalised neural noise in this study as increased trial-by-trial variability in the magnitude and/or the timing of responses to a discrete somatosensory stimulation event. Specifically, we examined trial-by-trial variability in responses to a single pulse of median nerve electrical stimulation [MNS]. Our results demonstrate that the P100 somatosensory evoked potential (SEP), which has been associated with conscious perception of tactile stimuli^11^, was significantly increased in the TS group. Importantly, however, while the timing, temporal variability, and spatial topography of early- and mid-latency SEP components (e.g., N20, P45, N60, P100) did not differ in the TS group, when compared to matched controls, trial-by-trial variability was substantially increased in the TS group, but this was normalised in response to stimulation. These findings may indicate that the trial-by-trial recruitment of neuronal sensorimotor populations is less stable at rest in individuals with TS compared to controls but may normalise in response to stimulation.

## Introduction

Tourette syndrome (TS) is a neurological disorder of childhood onset that is characterised by the occurrence of vocal and motor tics^1^. Tics are involuntary, rapid, abrupt, repetitive, recurrent, and non-rhythmic movements or vocalizations. TS or chronic tic disorder (CTD) affects ∼1-2% of children and adolescents and is the most common form of movement disorder in children^1^ . The pathophysiology of TS has been linked to dysfunction within cortical-striatal-thalamic-cortical (CSTC) brain circuits involved in the selection and initiation of movements^2.3^. Specifically, TS is associated with a reduction in the number of inhibitory fast-spiking parvalbumin interneurons in the striatum^2^, and it has been proposed that this might lead to increased neural ‘noise’ within the striatum and to downstream thalamo-cortical sensorimotor networks^3^.

Evidence in support of the proposal that the occurrence of tics may be due to increased neural noise resulting from striatal disinhibition has been obtained following the development of a convincing animal model of TS. Studies using this model have demonstrated that micro-injection of a gamma aminobutyric acid (GABA) antagonist into the striatum can reliably produce tic-like movements in non-human primates and rodents^4–8^. Thus, focal micro-injection of a GABA-A antagonist into the sensorimotor region of the striatum produces reversible increases in tic-like movements that are localised to a specific body part, commence within minutes of the injection, and typically last for 1 to 2 hours^6^. Furthermore, our unpublished studies using this animal model combined with electrophysiology and whole-brain single-photon emission computed tomography (SPECT) have demonstrated that striatal disinhibition, following micro-injections of the GABA-agonist picrotoxin delivered to the dorsal striatum in rodents, results in: tic-like movements; increased neural firing rates in the vicinity of the injection site; and more importantly, widespread increases in activation in remote brain areas distant from the site of injection^9^. While the concept of increased neural noise in TS is well-specified within the context of the striatal disinhibition model, its implications for behaviour more generally is not currently well understood.

A recent study used electroencephalography (EEG) to investigate whether so-called 1/f or scale-free noise was increased in individuals with TS^10^. 1/f noise can be calculated by first computing the power spectral density (PSD) of the EEG signals at different spectral frequencies (measured in Hz) and then by using linear regression to estimate the slope (ß) of the logarithm of the PSD with respect to frequency. Importantly, it is suggested that this 1/f noise slope estimate is proportional to the synchronised activity of neuronal populations^11,12^. Specifically, when neuronal firing is asynchronous, then the 1/f slopes are flatter than when neuronal firing is more synchronised. Thus, neural noise levels can be estimated from the steepness of the 1/f slope value, with steeper slopes indicating less noise and vice versa. Using a sensorimotor task, the authors of this study demonstrated that 1/f noise was indeed increased in individuals with TS compared to a matched group of neurologically healthy individuals. Importantly, however, in the context of the current study, the results from this study indicated that the difference in 1/f noise between the TS group and controls was only observed for higher (i.e., beta- and gamma-band) frequencies. The authors argued that this finding is likely due to neurobiological abnormalities associated with TS. This is likely to include alterations in GABA signalling^13^ which plays an essential role in the synchronisation of beta and gamma-band oscillatory brain activity^14^.

One likely consequence of increased neural noise is increased trial-by-trial variability in response to stimulation. High levels of neural variability (i.e., spontaneous levels of firing rate) are to be expected in the absence of stimulation, however it has been demonstrated that stimulus onset leads to a decline in neural variability even in circumstances that produce little or no change in the average firing rate. Churchland examined neural variability in 13 extracellularly recorded datasets from experimental animals and found that the decline in variability was observed for all types of stimuli tested, irrespective of whether the animal was awake, behaving or anaesthetized^15^. They propose that decline in variability is a general property of cortical responses, and that the cortical state is stabilized by an input. In the context of movement is has been proposed that decreases in trial-by-trial variability track the state of motor preparation as neural populations move toward an optimal response state^16,17^. Importantly, a study that investigated alterations in motor excitability that precede the execution of a voluntary movement in a group of individuals with TS reported that, unlike their age-matched controls, the TS group did not prior to the execution of volitional movements^18^.

To further investigate this issue we used EEG recording to examine how adults with TS responded to a single somatosensory stimuli (median nerve electrical stimulation [MNS]) compared to a group of matched neurologically healthy control participants. Importantly, for the purposes of this study we operationalised ‘neural noise’ as increased trial-by-trial variability in the magnitude and/or in the timing of responses to a discrete somatosensory stimulation event. Our predictions are as follows: 1. That for both groups, trial-by-trial variability will *decrease,* and inter-trial EEG phase coherence *increase*, in response to somatosensory stimulation 2. that individuals with TS will exhibit *increased* trial-by-trial variability in the magnitude and/or timing of EEG recorded events than controls. 3. That individuals with TS will exhibit *flatter* 1/f noise curves (i.e., log power spectral density as a function of spectral frequency) than controls at beta- and gamma-band frequencies. 3. That inter-trial EEG phase coherence estimates will be reduced in the TS group compared to matched healthy controls.

## Methods

### Participants

Data were collected from 20 right-handed participants, who had a confirmed diagnosis of Tourette syndrome (10F, mean age (± SD): 30.75 ±10.6). Prior the study, all the participants gave written informed consent and completed the following questionnaires: Edinburgh Handedness Inventory (EHI), Wechsler Abbreviated Scale of Intelligence (WASI), Premonitory Urge for Tics Scale – Revised (PUTS-R), Yale Global Tic Severity Scale (YGTSS). Further details can be found in Table 1. The experimental protocol was reviewed and approved by the local ethics committee (School of Psychology, University of Nottingham). Out of the 20 TS participants recruited for this study, one did not complete the EEG session due to discomfort.

**Table 1:** TS Participants Characteristics.

| ID | Age range | Sex | EHI | WASI | PUTS-R | YGTSS |
| --- | --- | --- | --- | --- | --- | --- |
| TS01 | 41-45 | F | R (90) | 109 | 38 | 73 |
| TS02 | 16-20 | F | R (90) | 100 | 72 | 81 |
| TS03 | 16-20 | M | R (85) | 119 | 63 | 27 |
| TS04 | 51-55 | M | R (100) | 124 | 39 | 32 |
| TS05 | 46-50 | M | R (95) | 125 | 31 | 31 |
| TS06 | 21-25 | F | R (100) | 119 | 61 | 47 |
| TS07 | 16-20 | F | R (100) | 108 | 20 | 45 |
| TS08 | 36-40 | F | R (55) | 109 | 45 | 41 |
| TS09 | 30-35 | M | R (90) | 125 | 54 | 47 |
| TS10 | 30-35 | M | R (100) | 106 | 42 | 43 |
| TS11 | 16-20 | F | R (100) | 97 | 59 | 56 |
| TS12 | 30-35 | M | R (85) | 128 | 19 | 14 |
| TS13 | 36-40 | M | R (65) | 80 | 41 | 71 |
| TS14 | 21-25 | F | R (85) | 117 | 32 | 57 |
| TS15 | 30-35 | M | R (100) | 111 | 44 | 74 |
| TS16 | 30-35 | M | R (100) | 126 | 48 | 45 |
| TS17 | 16-20 | F | R (75) | 115 | 16 | 34 |
| TS18 | 16-20 | F | R (80) | 90 | 80 | 74 |
| TS19 | 36-40 | F | R (40) | 99 | 60 | 50 |
| TS20 | 30-35 | F | R (100) | 100 | 24 | 69 |

EHI: Edinburgh Handedness Inventory. WASI: Wechsler Abbreviated Scale of Intelligence.
PUTS-R: Premonitory Urge for Tics Scale – Revised. YGTSS: Yale Global Tic Severity Scale

We also recruited 19 right-handed, neurologically healthy, participants as a healthy control group (HC). The HC group were age and sex matched to the TS group (10 F, mean age (± SD): 28.3, ± 6.7; t(29) < 1, p = .48). HC participants completed the EHI questionnaire and the informed consent forms.

### Electroencephalography (EEG) and Median Nerve Electrical Stimulation (MNS)

EEG measurements were obtained utilizing a 64-channel actiCHamp Plus system that incorporated active electrodes. EEG was recorded using Brain Vision Recorder software (Brain Products, UK) with a sampling frequency of the data was 25 kHz. Data was then down sampled to 500 Hz. The impedance of the electrodes was maintained below 30 μV for all the participants. The electrodes placed over the left and right mastoid (TP9 and TP10) were used as reference during these recordings.

Single pulse MNS was delivered over the dominant (right) wrist, with the electrodes placed over the median nerve (cathode proximal) using a Digitimer DS7A (Digitimer Ltd, UK) device. The stimulation pulse width was set at 0.2ms and the maximum voltage (Vmax) was set at 400 V. The motor threshold (MT) was measured individually for each participant and was defined as the minimum MNS intensity required to produce a visible thumb twitch (mean MT (± SD): TS group = 7.43 (1.84) mA; HC group = 7.03 (3.2) mA).

Participants were instructed to sit comfortably in a chair, resting their forearm on a desk placed in front of them. Each participant’s head was fixed on a chin rest to minimise head movements and participants were required to stare at a fixation cross presented on the wall, opposite to them. Before delivering MNS pulses to the hand, participants had the opportunity to ask questions and feel test pulses of MNS once the procedure had been explained to them.

In total, there were 75 MNS pulses delivered with a short break provided after each set of 25 pulses. To avoid anticipation effects, MNS was presented with a random inter-stimulus interval (ISI) of 11 ± 2 s. An in-house MATLAB script (MATLAB 2020a, Mathworks, Natick, MA) was used to control the timing and triggering of the MNS pulses. Whenever MNS pulses were triggered, markers were sent and saved in the EEG recordings via a trigger box (TriggerBox Plus, Brain Products, Gilching, Germany), which connected directly to the MNS device.

### Data analysis

EEG measurements were pre-possessed offline using BrainVision Analyzer 2 (Brain Product, GmbH). Prior to further analysis, electrodes exhibiting breakage or excessive noise were excluded following a visual inspection of the raw data.

The data were initially high-pass filtered at 0.5 Hz and low-pass filtered at 60 Hz using an 8-order Butterworth Infinite Impulse Response (IIR) and a 50 Hz notch filter to remove the electrical noise.

EEG recorded at the scalp may contain a mixture of signals arising from different sources, including electrical signals arising from muscle, or resulting from eye-blinks. This is a particularly important consideration to keep in mind when recording EEG from individuals with tic disorders like Tourette syndrome who may exhibit eye and facial motor tics. All EEG signals were inspected for evidence of eye-blinks or muscle artefact (see below). In addition, we conducted an Independent Component Analysis (ICA) of the EEG data to identify and remove muscle and ocular artefacts.

ICA is a method of blind source separation that separates a mixture of signals into a number of statistically independent components. It is based upon the assumption that the source signals are statistically independent of one another whereas the signal mixtures are not (Makeig et al., 1997). A common use for ICA is to identify and remove components associated with ocular and muscle artefacts. For the ICA, the reference electrodes were excluded, resulting in the segmentation of signals from the remaining 59 electrodes into epochs ranging from -300ms pre-stimulus to +600ms post-stimulus. This timeframe captures both baseline and post-stimulus neural activities, which are crucial for assessing stimulus-locked brain responses. ICA decomposes the multi-channel EEG data into independent components, which are then inspected for characteristics indicative of artefacts. Components reflecting muscle or ocular activity were identified based on their electrode locations, spectral properties, and scalp topographies. These artefact-related components were then excluded from further analysis. By removing these artifacts, ICA enhances the clarity of the neural signals, ensuring that subsequent analyses focus on the genuine sensory processes elicited by the stimuli.

Following ICA, the data were segmented into epochs based on the recorded MNS onsets, starting 200ms before the onset of MNS and ending 500ms after. Analyses focused on electrodes positioned over the primary somatosensory and motor cortical areas (Cz, CPz, C1, CP1, C3, CP3, FCz, FC1, FC3). Any epoch displaying abnormal noise levels or irregular trends in any electrode was excluded from further analysis. A semi-automatic inspection method was used to exclude any epochs with a signal amplitude ± 100 μV, maximal allowed voltage step of 50 μV/ms. Data were baseline corrected using the time window from -200 milliseconds to 0 milliseconds.

Following artefact rejection procedures, the somatosensory evoked potentials (SEP) were calculated as the average of the remaining trials. All participants had a minimum of 53 trials.

SEPs are typically recorded over the CP3 and C3 electrodes for right-handed participants and are characterised by well-defined onset components; the N1, which is a negative peak occurring ∼20ms post stimulation; N2 which is a negative peak occurring ∼60ms post stimulation; P2 which is a positive peak ∼100ms post stimulation, and the P3 which is a positive occurring peak 260ms post stimulus [**1**]. For brevity, only data recorded from CP3 will be reported here.

## Results

### Analysis of somatosensory evoked potential (SEP) component amplitude differences

Mean SEPs were computed for each individual for the time period spanning -200ms prior to the onset of MNS to 500ms. Note, however, that the focus in this study was primarily on examining the early (N1/P1) and mid (N2/P2) components of the SEP rather than late components. Grand averaged SEP waveforms for each group are presented in Figure 1A and the spatial topologies associated with the P1, N2, and P2 SEP components are presented in Figure 1B. Inspection of Figure 1A indicates that the SEP curves for the HC and TS groups are very similar following onset of the MNS. Importantly, there is no between-group differences in SEP amplitude or timing for the N1 (N20) or N2 (N60) SEP components. Inspection of Figure 1A does indicate however that the amplitude/timing of the P1 (P30) component (from +28-32ms) and the amplitude of the P2 (P100) from +92-108ms and the the P3 (P300) from +282-406ms components are substantially increased in the TS group between (Minimum effect size [Hedges G] > 0.5). The finding of an increased SEP P100 component in the TS group will be discussed below in the context of previous reports of sensory hypersensitivity in TS. Inspection of Figure 1B clearly indicates that the spatial topographies of the early P1, N2, P1 SEP components are highly similar for both groups.

**Figure 1.**
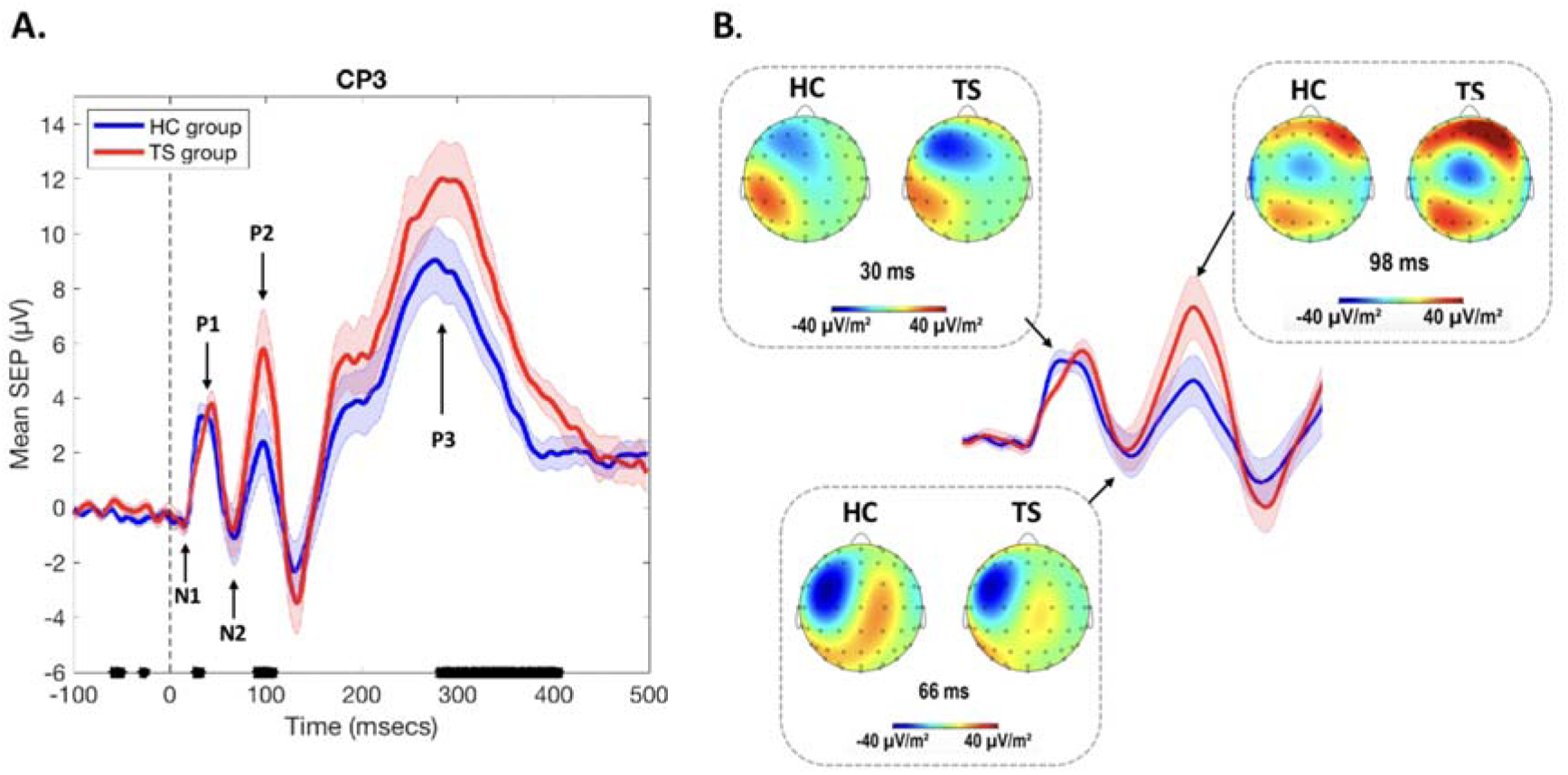
**A**. Grand average SEPs for HC and TS groups. Broken black vertical line indicates the onset of MNS. Black marks on x-axis reflect time points at which the curves for HC and TS groups were different (effect size [Hedges G] > 0.5). Shaded error bars are the standard error of the mean (SEM). **B**. Scalp topographies for HC and TS groups for P1 (30ms), N2 (66ms), and P2 (98ms).

### Analysis of SEP component timing differences

We hypothesised that increased neural noise in the TS group might result in alterations in the timing of SEP components, or more likely, in increased variability in the timing of SEP components. To examine this issue we conducted the following analyses. First, we computed a unified grand average SEP for the CP3 electrode upon the combined data from both groups. Then, based upon this unified grand average, we estimated the time of each of the SEP component peaks: N1 (+18ms), P1 (+40ms), N2 (+66ms), P2 (+98ms), and P3 (+272ms). Next, for each SEP component peak we determined a temporal window of a fixed size, defined *a priori*, that was centred at the mean time of each component. The sizes of these temporal windows were: N1 (± 10ms), P1 (± 20ms), N2 (± 20ms), P2 (± 20ms), P3 (± 50ms). Finally, the timing of the negative (N1, N2) peak value or positive (P1, P2, P3) peak value was determined on each trial for each individual. The results of this analysis are presented below (Table 2). Inspection of Table 2 shows that, as indicated in Figure 1A, the timing of the SEP P1 component was delayed by 1ms on average for the TS group. While this was a medium sized effect (-0.55), and statistically significant, the implications of a 1ms difference in the timing of the P1 remains to be determined. More importantly, there were no other between-group differences in the mean timing SEP components, and there was no evidence of increased *variability* in the timing of SEP components. In fact, the only significant between-group difference in mean variability was for the N2 component, where contrary to our predictions, the mean variability in timing was significantly *reduced* for the TS group. The absence of any between-group differences in the timing of SEP components will be discussed below.

**Table 2.** Mean times for SEP components for each group and mean variability (SD) in the timing of SEP components for each group.

| Mean time of SEP components (ms) |  |  |  |  |  |  |
| --- | --- | --- | --- | --- | --- | --- |
| Component | HC group |  | TS group |  | Effect size | p-value |
|  | Mean | SD | Mean | SD |  |  |
| N1 | 15.8 | 1.65 | 15.6 | 1.29 | 0.15 | 0.68 |
| P1 | 41.2 | 2.02 | 42.2 | 1.45 | **-0.55** | **0.05** |
| N2 | 64.9 | 2.89 | 65.5 | 2.84 | -0.2 | 0.27 |
| P2 | 96.9 | 3.35 | 97 | 3.28 | -0.31 | 0.17 |
| P3 | 284.3 | 6.65 | 285.1 | 6.38 | -0.13 | 0.34 |
| Mean variability (SD) in the time of SEP components (ms) |  |  |  |  |  |  |
| Component | HC group |  | TS group |  | Effect size | p-value |
|  | Mean | SD | Mean | SD |  |  |
| N1 | 5.6 | 0.66 | 5.7 | 0.73 | -0.1 | 0.39 |
| P1 | 5.7 | 0.56 | 6 | 0.79 | -0.41 | 0.11 |
| N2 | 10.1 | 1.59 | 9.3 | 1.18 | **0.64** | **0.02** |
| P2 | 10.4 | 1.7 | 9.9 | 1.18 | 0.36 | 0.14 |
| P3 | 25.8 | 3.39 | 26.1 | 2.09 | -0.1 | 0.38 |

### Analysis of variability of SEP amplitude

We predicted that increased neural noise in the TS group would result in an increase in trial-by-trial variability compared to healthy controls. To investigate this, we computed the standard deviation (SD) over all trials. for each individual, and for each time point. The mean SD values for each group are presented in Figure 2. Inspection of this figure (left panel) clearly shows that, as predicted, the TS group exhibit increased trial-by-trial variability compared to the HC group. Furthermore, this difference in variability is not confined to the period following stimulation onset, but instead is observed throughout the epoch (Hedges G > 0.5). Inspection of Figure 2 also clearly indicates that mean variability increases in both groups in response to the onset of stimulation, and the rate of increase and the timing of the asymptote looks to be highly similar for both groups. To examine this further, we re-analysed the SD data having first standardised (z-score) the data for each trial. This has the effect of removing any baseline differences in magnitude. Mean standardised SD data for each group is shown in the left panel of Figure 2. Inspection of this figure indicates that the rate of increase in variability after stimulation onset, and the asymptote, is almost identical for the HC and TS groups.

**Figure 2.**
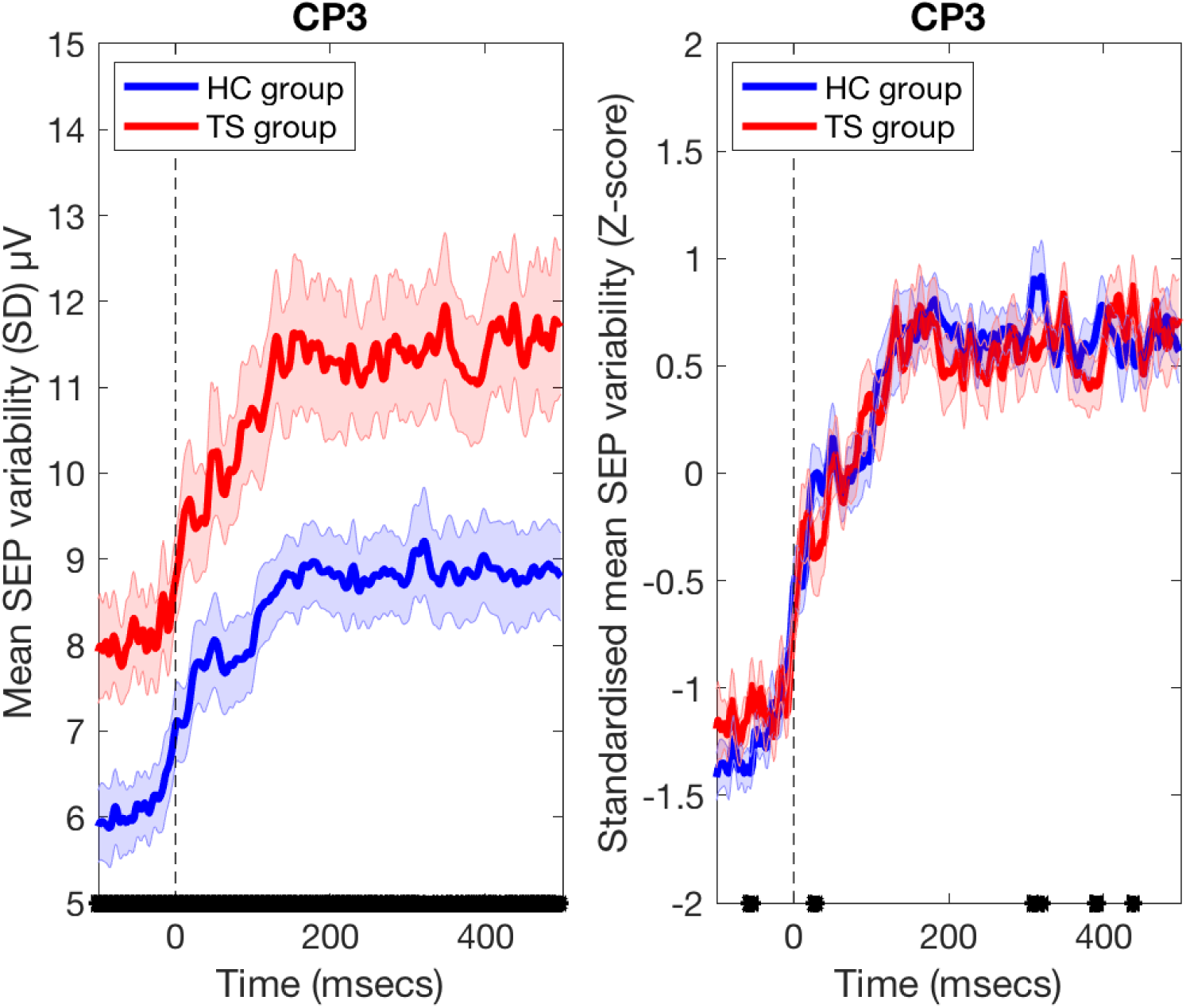
Left Panel. Mean MEP variability (SD) for HC and TS groups. Broken black vertical line indicates the onset of MNS. Black marks on x-axis reflect periods where curves for HC and TS groups differ (effect size [Hedges G] > 0.5). Right Panel: Shows the mean standardised MEP variability (z-score) for HC and TS groups. This illustrates that the increase in average variability (SD) increases at the same rate after stimulation onset in both groups. Shaded error bars are SEMs.

### Analysis of Coefficient of Variation (CV)

Previous studies have demonstrated that a metric measuring trial-by-trial variability, the Fano factor (the variance in firing rate divided by the mean firing rate), decreases following stimulation onset, and that a reduction in variability may be a general property of cortical responses to stimulation, and reflects stabilisation of the cortical response to an input.^15^. To further investigate any between-group differences in trial-by-trial variability, we computed a comparable metric, the coefficient of variation (CV) over all trials. for each individual, and for each time point. The CV standardises the SD with respect to the mean value and is computed by taking the SD/mean ratio. To examine this, the CP3 EEG data for the period from 200ms before stimulation onset to 200ms after stimulation onset was separated into four 100ms epochs: -200 to -101ms, -100 to -1ms, 0 to 99ms, 100 to 200ms and the median CV (an index of trial-by-trial variability) was calculated for each epoch and for each individual. Data were then analysed using a 2-way mixed ANOVA with the between-subject factor Group and the Within-subject factor Epoch. Relevant means are presented in Figure 3.

**Figure 3.**
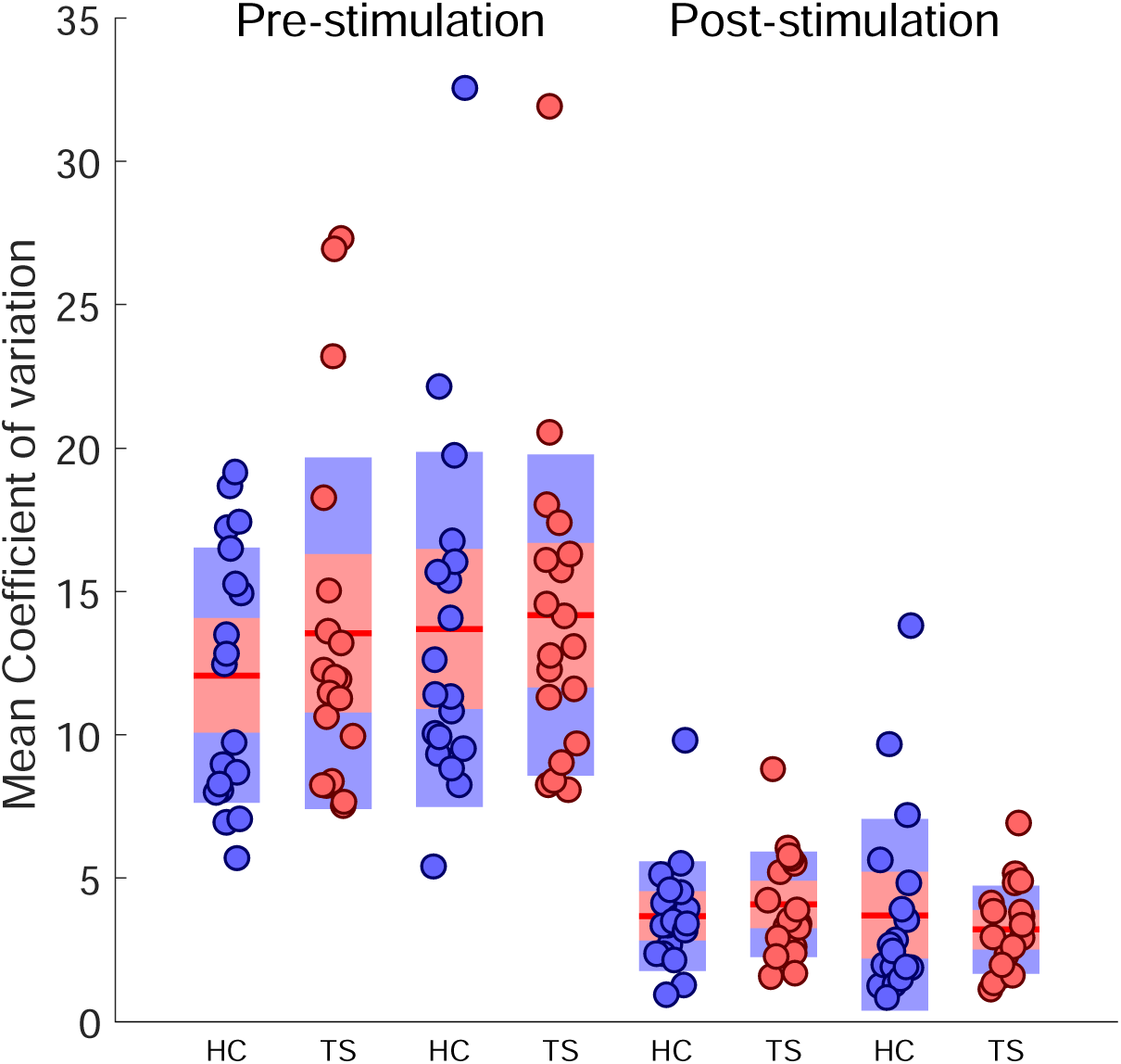
Mean coefficient of variation (CV) values for pre-stimulation period (-200 to -1ms) compared to post-stimulation (0 to 199ms) period. Both groups show the predicted substantial reduction in CV following stimulation onset. Importantly, there are no between-group differences in CV either before or after stimulation onset.

The ANOVA revealed no main effect of Group (F(1,36) < 1, p = .55) and no Group x Epoch interaction effect (F(3,108) < 1, p = .78). By contrast, the ANOVA revealed a significant main effect of Epoch (F(3,108) = 71.4, p < .00001). Inspection of Figure 3 clearly shows that this effect is due to mean CV values decreasing substantially for both groups in the epochs following the onset of stimulation.

### Power Spectral Density (PSD) analyses

As noted above, aperiodic (1/f) noise can be estimated from the slope of the logarithm of the PSD of the EEG with respect to frequency, and it is suggested that this 1/f noise slope estimate is proportional to the degree of synchronised neural activity^11,12^. Based upon a previous study^10^, we predicted that the mean 1/f slopes would be flatter for the TS group (indicating increased neural noise) in comparison with healthy controls, particularly for frequencies within the beta (13-30Hz) and gamma (> 30Hz) bands. To examine this we used Welch’s averaged, modified periodogram method to calculate PSD within the frequency range 1-45Hz for each individual for all trials. Mean log PSD values with respect to frequency are presented for each group in Figure 4A. Inspection of this figure clearly shows that mean spectral power is increased for TS group compared to HC group (minimum Hedges G > 0.5) at the high-beta (>25Hz) and gamma (>30Hz) band frequencies and that, as predicted, the gradient for the TS group appears flatter as consequence. To examine this, we followed the approach used previously^10^ by fitting for each individual, a linear regression for log PSD with respect frequency. The mean fitted curves for each group are presented in Figure 4B and confirm that the slope of the mean linear fit for TS group is indeed flatter than that for the HC group, and that the two curves differ from one another at beta and gamma-band frequencies (minimum Hedges G > 0.5).

**Figure 4.**
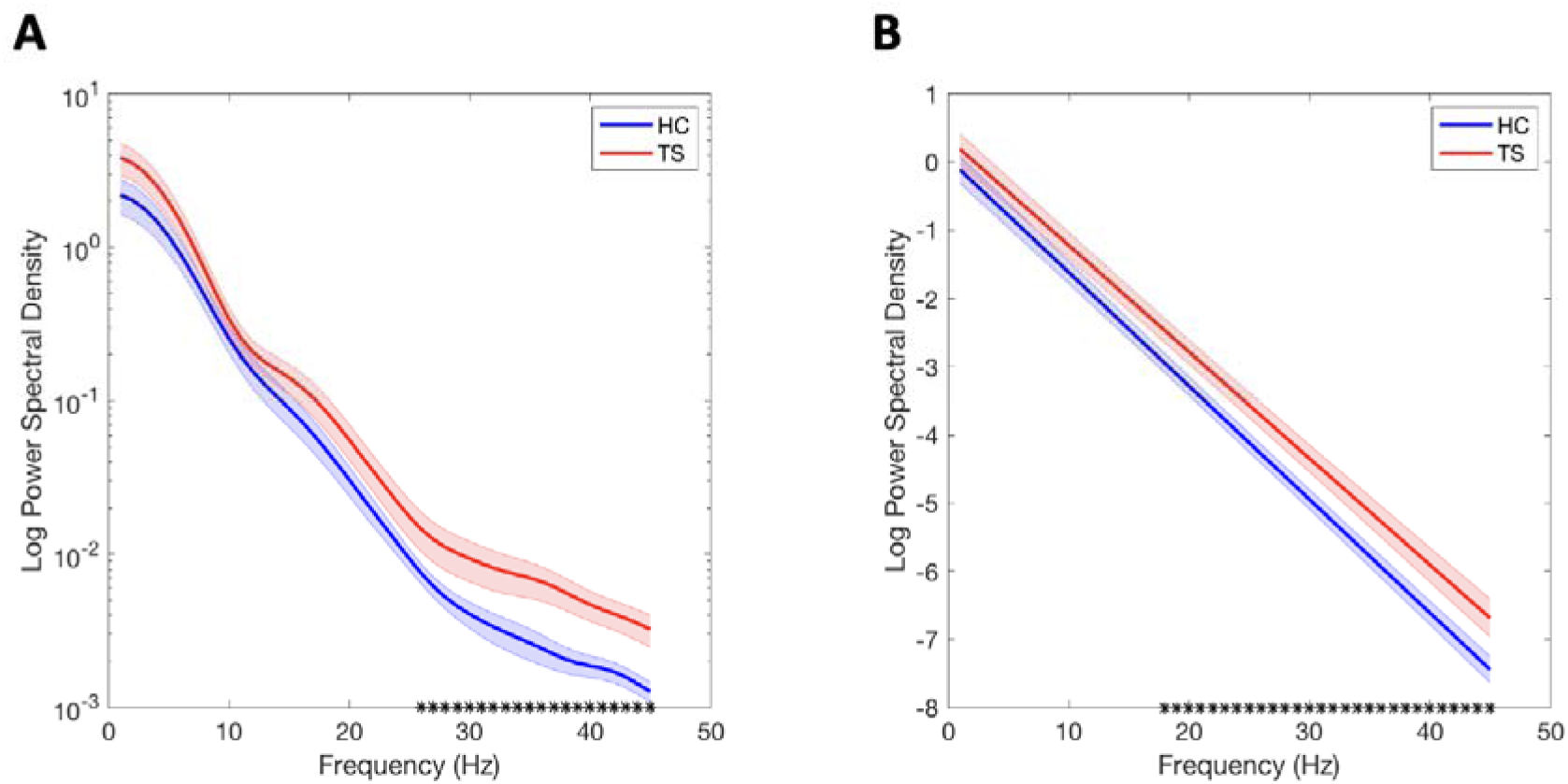
**A**. Power Spectral Density (PSD) estimates were calculated for each participant using Welch’s averaged, modified periodogram method for frequencies from 1-45Hz. These data were then averaged to provide mean PSD estimates. Black symbols on the x-axis reflect frequencies at which mean PSD differed between groups (Effect size [Hedges G] > 0.5). Shaded error bars indicate the SEM. Linear regression was used to obtain linear fits of the logarithm of the PSD with respect to frequency for each individual. **B**. shows the mean linear fits for each group. Shaded error bars indicate the SEM. Black symbols on the x-axis reflect frequencies at which mean PSD differed between groups (Effect size [Hedges G] > 0.5).

### Analysis of inter-trial phase coherence

It has been demonstrated previously that decreases in trial-by-trial variability track the state of motor preparation as neural populations move toward an optimal response state^16,17^. Furthermore, it is likely that this progression toward an optimal response state would be reflected in an increase in the synchronous activity of the activated neural population. Based upon this assumption we predict that inter-trial phase coherence would increase in response to stimulation as a consequence of increased neural synchronisation. To examine this, we used the EEGLAB *phasecoher* function to compute mean inter-trial phase coherence estimates for alpha-band (8-12Hz), beta-band (13-30Hz) and low gamma-band (31-45Hz) frequencies for each individual and for each group. Relevant data are shown in Figure 5. Inspection of Figure 5 suggests that, as predicted, inter-trial phase coherence increases in response to somatosensory stimulation. To examine this, we computed for each participant, and for each frequency band (i.e., alpha, beta, and low gamma), the mean phase coherence for the 50ms period immediately preceding stimulation onset and the 100ms period immediately following stimulation onset. These data were then compared using separate 2-way mixed ANOVAs with the between-subject factor of Group and the within-subject factor Period (pre vs. post).

**Figure 5.**
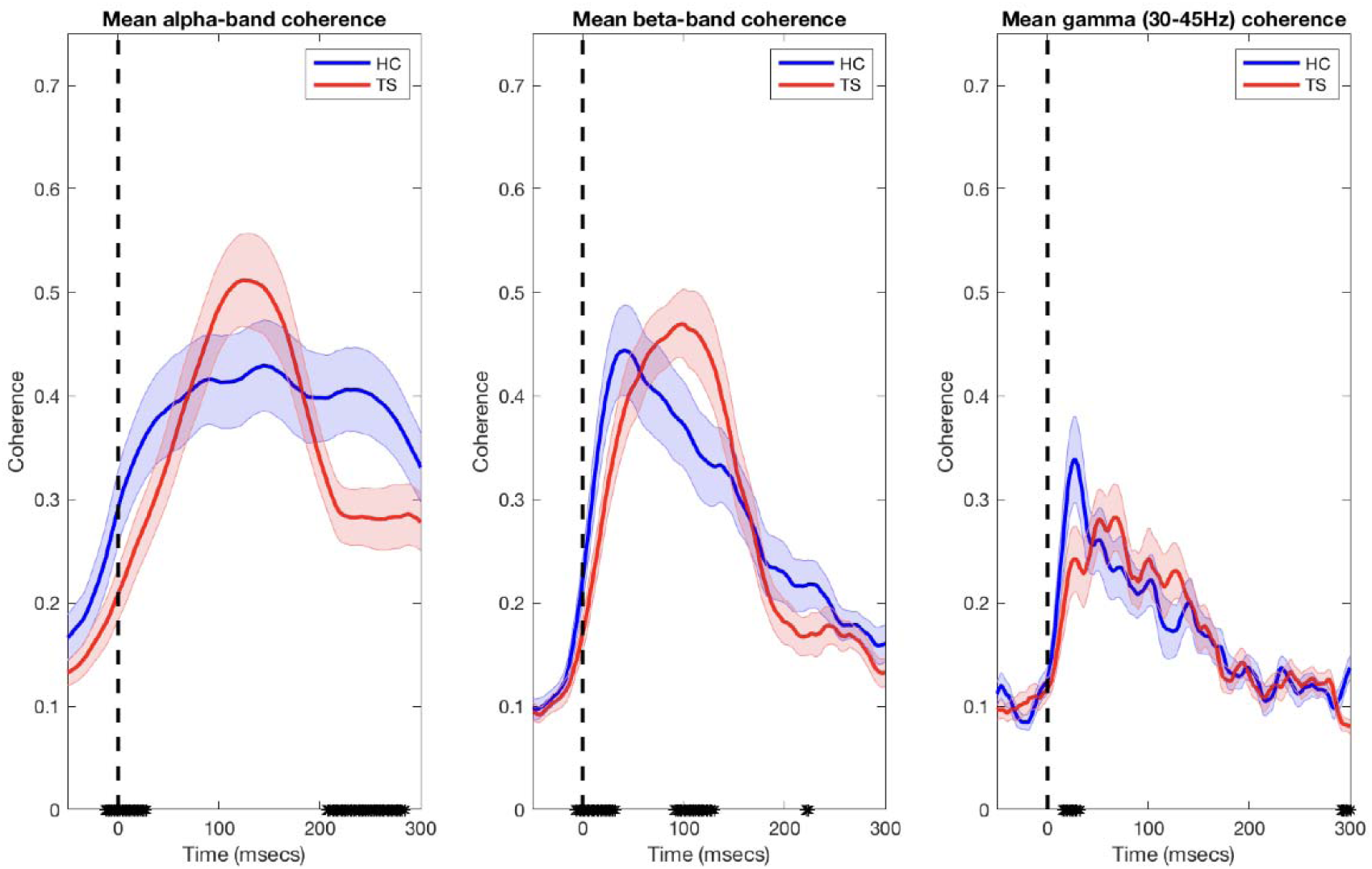
Mean inter-trial phase coherence estimates for theta, alpha, beta and low-gamma band frequencies computed for each group. In all cases, mean inter-trial phase coherence increased after stimulation onset (indicated by the broken black vertical lines). This increase in phase-coherence was modest for theta-band frequencies but more pronounced for alpha, beta and low-gamma frequencies. The black symbols on the x-axes indicate time periods at which the mean coherence estimates for each group differed (effect size [Hedges G] > 0.5).

The ANOVA for alpha-band (8-12Hz) frequency coherence revealed no significant main effect of Group (F(1,36) < 1, p = .87) or a Group x Period interaction (F(3,108) < 1, p = .67), but a significant main effect of Period (F(3,108) = 58.8, p < .00001). This is explained by mean inter-trial alpha phase coherence increasing substantially immediately following the onset of stimulation.

The ANOVA for beta-band (13-30Hz) frequency coherence also revealed no significant main effect of Group (F(1,36) < 1, p = .91) or a Group x Period interaction (F(3,108) < 1, p = .66), but a significant main effect of Period (F(3,108) = 59.8, p < .00001). Again, this is explained by mean inter-trial beta phase coherence increasing substantially immediately following the onset of stimulation.

Finally, stimulation. the ANOVA for low gamma-band (31-45Hz) frequency coherence also revealed no significant main effect of Group (F(1,36) < 1, p = .93) or a Group x Period interaction (F(3,108) < 1, p = .66), but a significant main effect of Period (F(3,108) = 63.1, p < .00001). This effect is also explained by mean inter-trial gamma phase coherence increasing substantially immediately following the onset of stimulation.

We hypothesised that increased phase coherence following the onset of stimulation would likely reflect an increase in the synchronous activity of the activated neural population as it progressed toward an optimal response state. While a substantial increase in phase coherence is demonstrated for both groups, inspection of Figure 5 illustrates that there are between-group differences in mean phase coherence, which for the most part reflect reduced phase coherence in the TS group. Specifically, for alpha, beta and gamma-band frequencies, the early rate of rise in phase coherence (∼0-40ms) is *shallower* in the TS group compared to the healthy control group (minimum Hedges G > 0.5). This finding is consistent with the proposal that increased neural noise in the TS group may result in a delay neural synchronisation.

For alpha-band frequencies, the mean phase coherence is sustained at a high level for *longer* in the HC group compared to the TS group (minimum Hedges G > 0.5). Finally, for beta-band frequencies, mean phase coherence is *increased* in the TS group relative to controls for a period ∼100ms after stimulation onset (minimum Hedges G > 0.5). Interestingly, this coincides with the timing of the SEP P2 (P100) component, the magnitude of which was also significantly increased in TS group. This effect will be discussed below.

### Association between key dependent measures and clinical scores

To examine whether the key dependent measures reported above were associated with biographic and clinical data from the TS group, we conducted a series of multiple linear regression analyses with the following variables as predictor variables: Age, Sex, WASI score, PUTS-R score, YGTSS-motor score, YGTSS-vocal score, and YGTSS-impairment score. The results of these analyses are presented in Table 3. Note, Age, Sex, and YGTSS-vocal score were not statistically significant predictors in any of the analyses reported below.

**Table 3.** Results of multiple linear regression analyses.

| Dependent Variable | Predictor | r-value | R-square | F-value | p-value |
| --- | --- | --- | --- | --- | --- |
| SEP P2 amplitude | YGTSS-impairment score | 0.54 | 0.27 | 6.8 | 0.02 |
| Pre-post stimulation alpha coherence | YGTSS-motor score | -0.53 | 0.28 | 6.7 | 0.02 |
| Pre-post stimulation gamma coherence | PUTS-R score | 0.45 | 0.2 | 4.3 | 0.05 |
| Pre-post stimulation CV | PUTS-R score | 0.48 | 0.23 | 5.1 | 0.04 |
| Mean SEP variability (SD) | WASI score | -0.59 | 0.35 | 9.1 | 0.008 |
| PSD (25 - 45 Hz) | WASI score | -0.63 | 0.4 | 11.4 | 0.004 |

#### Magnitude of SEP P2 (P100) component

The multiple regression analysis revealed that P2 magnitude is predicted by tic severity (YGTSS-impairment score: r = 0.54, R^2^ = 0.27, F=6.8, p = 0.02). Larger P2 SEP values are associated with increased tic impairment. This finding will be discussed below in the context of sensory hypersensitivity in TS. No other variables were significant predictors.

#### Change in alpha-band coherence

Mean alpha-band coherence was calculated for the 100ms period immediately following stimulation onset and subtracted from the 50ms period immediately preceding stimulation onset. The multiple regression analysis revealed that change in alpha coherence is predicted by tic severity (YGTSS-motor score: r = -0.53, R^2^ = 0.28, F=6.7, p = 0.02). Increased severity of motor tics was associated with larger increases in alpha-band coherence following somatosensory stimulation. This finding will also be discussed below in the context of sensory hypersensitivity in TS. No other variables were significant predictors.

#### Change in gamma-band coherence

Mean gamma-band coherence was calculated for the 100ms period immediately following stimulation onset and subtracted from the 50ms period immediately preceding stimulation onset. The multiple regression analysis revealed that change in gamma coherence is predicted by premonitory urge severity (PUTS-R score: r = 0.45, R^2^ = 0.2, F=4.3, p = 0.05). Individuals with higher premonitory urge scores exhibited the lowest increases in gamma-band activity following stimulation onset. No other variables were significant predictors.

#### Change in coefficient of variation (CV)

Mean CV values were calculated for the 200ms period immediately following stimulation onset and subtracted from the 200ms period immediately preceding stimulation onset. The multiple regression analysis revealed that change in CV is predicted by premonitory urge severity (PUTS-R score: r = 0.48, R^2^ = 0.23, F=5.1, p = 0.04). Individuals with higher premonitory urge scores exhibited the greatest decrease in variability (CV) following stimulation onset. No other variables were significant predictors.

#### Mean SEP variability (SD)

Mean SD values were calculated for the entire epoch. The multiple regression analysis revealed that mean SD values are predicted by IQ score (WASI: r = -0.59, R^2^ = 0.35, F=9.1, p = 0.008). No other variables were significant predictors.

#### Mean Power Spectral Density (PSD)

Mean PSD values were calculated for each individual for 25-45Hz frequencies. The multiple regression analysis revealed that mean PSD values are predicted by IQ score (WASI: r = -0.63, R^2^ = 0.4, F=11.4, p = 0.004). No other variables were significant predictors.

## Discussion

This study investigated the proposal that the occurrence of tics in Tourette syndrome results from increased sensorimotor noise. To investigate this, we operationalised increased sensorimotor noise as an increase in trial-by-trial variability in EEG response (measured at the CP3 electrode) to a single somatosensory stimulus, specifically, a single pulse of electrical stimulation to the median nerve. The key findings of this study are discussed below.

### No difference in amplitude, timing and topography of early and mid-latency SEP components

Our results demonstrated that, for the most part, the mean amplitude, timing and spatial topography of early and mid-latency components of the somatosensory evoked potential (SEP) was not different in the TS group compared to healthy controls. Furthermore, and contrary to our predictions, there was no evidence that the timing of SEP components was more variable in the TS group than that observed for the control group. This is important to keep in mind when considering the finding of increased trial-by-trial variability in the SEP observed in the TS group.

Two discrete between-group differences in SEP amplitude/timing were observed: a 1ms slowing in the TS group in the timing of the P1 SEP; and a substantial increase in the magnitude of the P2 (P100) SEP. We believe that the former is a trivial and likely unreliable finding, and for this reason it is not discussed further. By contrast, we believe that the finding of an increased P100 SEP component in the TS group is important and is discussed below.

### Increased P100 and sensory hyper-sensitivity in TS

There is mounting evidence to indicate that the majority of individuals with TS experience tactile hypersensitivity – i.e., a relatively constant heightened awareness of tactile stimuli^19,20^. However, psychophysical investigation of detection thresholds for tactile stimuli has revealed no differences between individuals with TS and matched healthy controls^19^ and this finding has been interpreted as evidence that tactile hyper-sensitivity is most likely related to altered central processing associated with the stimulus evaluation, rather than any alterations in initial somatosensory processing. Furthermore, individuals with TS do not appear to habituate normally to repetitive stimuli, suggesting an inability to effectively filter redundant sensory input^19,20^.

Importantly, the P100 component of the SEP has previously been implicated in the conscious perception and evaluation of tactile stimuli^21–23^. In the current study we observed that the P100 SEP component was significantly increased in the TS group compared to healthy controls, and that the magnitude of this P100 component was significantly correlated with tic severity (i.e. increased P100 amplitude was positively associated with increased tic severity (YGTSS-impairment). Furthermore, while for the most part, the increase in inter-trial phase coherence that occurred following the onset of somatosensory stimulation was significantly *reduced* in the TS group, this was not the case for beta-band coherence, which was demonstrated to be significantly *increased* in the TS group, relative to controls, but only for the period coinciding with the timing of the P100 SEP component. These findings are consistent with the proposal that central processing associated with somatosensory stimulus evaluation is altered in individuals in TS leading to heightened awareness of tactile stimuli.

### Increased trial-by-trial variability of SEPs in Tourette syndrome

In the current study we assumed that increased neural noise in TS would manifest as increased trial-by-trial variability compared to that observed for healthy controls. We also assumed that neural noise would reduce when neural firing is synchronous rather than asynchronous and as neural populations move toward a synchronised response to stimulation^16,17^. These assumptions were examined in several ways.

We initially examined trial-by-trial variability (which we operationalised as the standard deviation of individual SEPs) and demonstrated that trial-by-trial variability was, as predicted, substantially larger in the TS group compared to controls. However, this increased variability in the TS group was observed throughout the trial and was not limited solely to the period following median nerve stimulation. I should be noted that a comparable finding was previously reported in individuals with autism spectrum disorder (ASD)^24^; where it was concluded that individuals with ASD were less able to synchronise neural activity compared to neurotypical individuals. Here we suggest that trial-by-trial recruitment of neuronal sensorimotor populations may less stable at rest in individuals with TS compared to neurotypical controls. This increased variability at rest is likely due to abnormal subcortical activity, specifically, dysfunctional striatal-thalamic efferents that target cortical sensorimotor areas^2^, that may normalise in response to somatosensory stimulation (see below).

As noted above, one way of examining neural noise is to estimate the 1/f slope following a Power Spectral Density (PSD) analysis. Specifically, it is proposed that the 1/f slope estimate is proportional to the synchronised activity of neuronal populations, with flatter 1/f slopes associated with more *asynchronous* neural firing ^11,12^. In the current study we demonstrated that mean PSD was increased in the TS group at higher frequencies (i.e., high-beta (>25Hz) and gamma (>30Hz) band frequencies). As a result, the gradient of the estimated 1/f slope was flatter for the TS group compared to controls. This finding is consistent with the result of a previous study that reported a flatter 1/f slope in TS^10^.

Importantly, it has been proposed that GABA-mediated neural transmission plays a key role in promoting synchronised activity in brain networks^26,27^, and that dysfunction in GABAergic inhibition leads to abnormalities in brain oscillations in several brain disorders^26^. We suggest that the current finding of increased trial-by-trial variability in TS is likely linked to altered GABAergic inhibition in TS^2,3^. Particularly, reduced GABA-A receptor mediated inhibition within cortical sensorimotor areas previously reported in TS^25,28^.

To further investigate group differences in variability, we also examined standardised coefficient-of-variation [CV] (i.e., the standard deviation/mean ratio) values for each group, including how CV altered following stimulation. Previous studies have demonstrated that the Fano factor (i.e., the variance of the neural firing rate divided by the mean firing rate) decreases in response to stimulation^15^, and it is proposed that a reduction in variability may be a general property of cortical responses to stimulation that reflects stabilisation of the cortical response to input^15^. Our results demonstrated that mean CV values decreased substantially for both groups in response to stimulation. Importantly, the changes in CV values were comparable for both groups.

Decreases in trial-by-trial variability have been shown to track the state of motor preparation as neural populations move toward an optimal response state^16,17^, and to decrease in response to stimulation^15^. We hypothesised that this progression toward an optimal response state would be reflected by an increase in the synchronous activity of the activated neural population and predicted that inter-trial phase coherence would therefore increase in response to somatosensory stimulation.

Our findings are broadly consistent with the proposal that increased neural noise in the TS group results in an altered rate of neural synchronisation and/or delayed neural synchronisation. Consistent with this proposal, mean Mu/Alpha-band phase coherence was demonstrated to be sustained at a higher level for *longer* in the control group compared to the TS group. However, the interpretation of any observation of increased/decreased coherence in the TS group must be consider the presumed functional role of increased/decreased neural synchronisation. Thus, for beta-band frequencies, by contrast, we observed that mean phase coherence was demonstrated to *increase* in the TS group, relative to controls, for the period that coincided with the timing of the increased SEP P2 (P100) component observed for TS group. In this case, it is likely that this increased beta-band phase coherence reflects the increased P2 (P100) SEP response to somatosensory stimulation that is likely linked to increased central processing of somatosensory stimuli associated with heightened awareness of tactile stimuli in TS.

Taken together, these results suggest that trial-by-trial recruitment of neuronal sensorimotor populations may less stable at rest in individuals with TS compared to neurotypical controls. This increased variability at rest in the TS group is likely due to abnormal subcortical activity, specifically, dysfunctional striatal-thalamic efferents that target cortical sensorimotor areas^2^. Variability reduced substantially in both groups in response to stimulation. This is consistent with the proposal that progression toward an optimal response state will result in increased synchronous activity of the activated neural population. In this context it is important to note that the rate at which variability reduces was comparable in both groups. This finding is consistent with a previous report showing that abnormal cortical activity at rest in TS normalises during motor performance^25^.

## Conclusion

In the current study, we used electroencephalography (EEG) to investigate increased neural noise in a group of 19 adults with TS compared to a matched neurotypical control group. We operationalised neural noise in this study as increased trial-by-trial variability in the magnitude and/or the timing of responses to a single pulse of median nerve electrical stimulation [MNS]. We demonstrated that, while the P2 (P100) SEP (previously associated with conscious perception of tactile stimuli) was significantly increased in the TS group, the timing, temporal variability, and spatial topography of early- and mid-latency SEP components (e.g., N20, P45, N60, P100) did not differ in the TS group, when compared to matched controls. However, trial-by-trial variability was substantially increased in the TS group, but this was shown to normalise in response to stimulation. Together these findings suggest that the trial-by-trial recruitment of neuronal sensorimotor populations is less stable at rest in individuals with TS compared to controls but may normalise in response to stimulation.

## Data Availability

All data produced in the present study are available upon reasonable request to the authors

## Acknowledgements

Stephen Jackson and Aikaterini Gialopsou were supported by research grants from Medical Research Council (T032588) and Parkinson’s UK. Stephen Jackson and Mairi Houlgreave were supported by the NIHR-funded Nottingham Biomedical Research Centre. Stephen Jackson was also supported by an EPSRC IAA grant. We are grateful to Matt Brookes for helpful discussions.

